# Adherence to the 2016 PAHO Clinical Practice Guideline on Dengue in a Hospital in Northern Peru, 2022-2023

**DOI:** 10.1101/2024.07.25.24310969

**Authors:** Franco E. León Jiménez, Nataly BF. Mendoza-Farro, Adriana Montoya Reategui, Joel Emmanuel Inga-Chero, Karim Dioses Diaz, Moisés Barranzuela-Herrera, Sophia Cavalcanti Ramírez, Luz M. Moyano, de Neuroepidemiology and Science of life working group

**Affiliations:** School of medicine, Universidad Cesar Vallejo, Perú (FLJ, UCV-Trujillo; KDD, LMMV UCV-Piura); Department of Medicine, Hospital de la Amistad, Peru Korea Santa Rosa II-2, Piura-Perú (AMR, SCR Infectology service, MBH, medical resident, FLJ, KDD medicine service); Epidemiology and environmental health unit, Hospital de la Amistad, Perú Korea Santa Rosa II-2, Piura, Perú; School of Medicine, Universidad Nacional de Piura, Perú; Scientific society of medical students, Universidad Nacional de Piura, Perú

**Author notes:** **Contact email:** Corresponding author, Luz María Moyano Vidal.

**Keywords:** Dengue, Adherence, Clinical Practice Guidelines, Mortality

## Abstract

**Background:** Effective dengue management enhances the chances of survival. The level of adherence to the suggestions in northern Peru is uncertain. The primary aim of the study was to assess adherence to the 2016 PAHO guideline on dengue in a hospital located in northern Peru during the period from 2022 to 2023.

**Methodology/principal findings:** The study performed a cross-sectional design and exploratory analysis, reviewing 141 medical records. Fifty-four percent were from 2023; 65.9% were from women; 46.1% came from another healthcare center; 76.6% had diagnosis of dengue with warning signs; 20.7% had severe dengue; and 18.44% died. We found at least one error in the classification of severity and/or treatment in non-hospital healthcare facilities (91.5%), triage (88.5%), and uviclin/observation (52.6%). Errors in classification and inadequate hydration in the non-hospital healthcare centers, triage, and uviclin/observation were: 35.8%/53.7%, 30.6%/76.6%, and 10.1%/45.6%, respectively. Persistent errors were inadequate hydration in triage (76.86%) and urinary flow in the center (73.13%). In a bivariate analysis, mortality was associated with older age (p = 0.035), having a case from 2023 (p = 0.0073), having a case from the study hospital (p = 0.083), and having severe dengue (p<0.001). In the multivariate analysis, only severe dengue (OR 318.4, 95% IC [33.8-2996], p<0.001) was associated with mortality.

**Conclusions/significance:** We found a high frequency of misclassification and management errors in these three scenarios, but they were not associated with higher mortality.

**Author summary:** According to the PAHO 2016 Dengue Guidelines, this descriptive study found deficiencies in the diagnosis of severity and treatment in a public hospital of the Ministry of Health of Peru, located in the region with the highest dengue incidence rate in 2023. We identified these deficiencies in the pre-hospital phase, as well as in the emergency, observation, and dengue management units, which had a high mortality rate of 18%. While it is true that management errors were not associated with higher mortality in the exploratory analysis, study biases may explain this result. We must take these findings into account when investigating the effectiveness of the health worker training that took place between 2023 and 2024, employing a master-class methodology and awaiting a before and after analysis to gauge its impact on knowledge. The need for improvements in health data management highlights the need to digitize information and electronic health records.

## Introduction

In the Americas, in 2023, there were 4’563, 485 cases of dengue, 7,653 cases of severe dengue (0.16%) and 2,339 deaths (0.05% mortality). After Brazil, Peru had the second highest number of cases (274,277 cases), with 441 deaths (0.16%), which is 3.2 times the average for the Americas (1). Peru recorded a total of 13,010 cases and 8 fatalities up until epidemiological week 5. Among these instances, the city of Piura in the northern region reported 2,153 cases. (2).

Although this virus’s mortality is less than 1%, complications and deficiencies in early management can increase it. The most key factors for improving prognosis are early syndromic diagnosis, patient individualization, monitoring, and adequate hydration. (3).

In 2016, the Pan American Health Organization (PAHO) developed the *Guidelines for patient care in the Region of the Americas 2nd edition,* based on available evidence from cohort studies and expert opinions. in which they emphasize the correct initial management according to risk factors and symptomatology for adequate triage and treatment, according to the widely recognized classification: Dengue without warning signs (DWWS), Dengue with warning signs (DWS) and Severe Dengue (SD) (4). Within the Peruvian contingency plan of the Ministry of Health (MINSA) against Dengue, in October 2023, a National Directive was issued ensuring a budget of 107,585,222 PEN for the program *017 Metaxenic and zoonotic diseases*, of which more than 16,000,000 PEN were assigned to the MINSA and more than 91,000,000 PEN to the Regional Governments, aimed to strengthen the skills and competencies of healthcare professionals in the management and approach of this problem and to develop research in this pathology. Likewise, this regulation exposed the deficit in adherence to the Clinical Practice Guidance (CPG) recommendations and the lack of research and exploration of the problems related to it (5). On the other hand, in our country, in March 2024, the Technical Health Standard No. 211-MINSA/DGIESP-2024 was implemented, with clear specifications on prevention and treatment at diverse levels of care (6). This allowed us to adopt an anticipatory and prospective attitude in the evaluation of the patient. To implement a guide, it is necessary to educate in all areas of care and train all healthcare personnel through validated educational interventions with theoretical and practical content. In other words, it is necessary to train and measure to know how we are, where we are going, what we need to improve and how we will do it. Adherence to recommendations can provide better clinical outcomes (8) (9).

There is evidence that the knowledge, attitudes, and practices in the comprehensive management of this endemic disease are far from ideal. In the 2022 outbreak, studies on patients hospitalized for DWS and SD in a Level II-2 hospital of the Ministry of Health of Piura found delays in reference time, prolonged time in triage and frequent errors in severity classification, hydration, and monitoring (10). In Colombia, a review of 43 medical records for the characterization of adherence to the 2010 PAHO CPG, found that the overall adherence level was 67.4% and the items with the lowest compliance were: evaluation capillary refill, search for hemorrhages in the skin and mucous membranes, evaluation of warning signs and examination of the central nervous system (11). In another Colombian study in pediatric patients, it was found that 77.18% were correctly classified and 28.9% received adequate hydration management (12).

Since Piura is the city with the most cases of dengue reported in the period 2023–2024 and deficiencies have been evident in 2022, this study aims: a) to describe the most frequent errors in the classification of the severity and management of admitted dengue cases to a public hospital in the city of Piura in the years 2022–2023 and b) to explore their association with the severity and mortality of dengue.

## Material and methods

### Design

This is a cross-sectional descriptive study with exploratory analytical analysis.

### Study site

The region of Piura (2,127,000 inhabitants) is the second department with the largest population in Peru (13). The Ministry of Health of Peru rates Hospital de la Amistad, Peru-Korea Santa Rosa, the study’s site, as Level II-2. (14) In 2023, it had Emergency Room Triage (20 beds), Pediatric Emergency Services (12 beds), Obstetrics and Gynecology Emergency Services (10 beds), Medicine Emergency (20 beds), and Surgery Emergency (14 beds); two Dengue Monitoring Units named Uviclin (Uviclin 1: [12 beds] and Uviclin 2: [40 beds]); an Intensive Care Unit (pediatrics, intermediate care, and general: [32 beds]); Internal Medicine Hospitalization (50 beds); Surgery (40 beds); Obstetrics and Gynecology Hospitalization (14 beds); and Pediatrics (13 beds). Up until the last patient’s description, the hospital had a total of 277 beds available. The hospital has handwritten registration and information management does not have electronic medical records for hospitalization.

### Population and study sample

The study population was the medical records of patients with DWS and SD, hospitalized in at least one of the aforementioned services. Medical records from 2022 and epidemiological weeks 18-25 of 2023 (peak of the outbreak) were included. Sampling was by convenience according to the availability of the medical records. With a population of 939 records, confidence level of 97%, design effect of 1 and frequency of non-registration/no data available for mean arterial pressure in Triage of 11.3% (9) (previous study from 2022), the calculated sample size was 158 stories. The records of patients with or without referral from another health care facilities and followed until their permanence in observation and/or Uviclin and/or ICU were considered.

### Information from medical records and definitions

SCR, AMR (Infectious diseases physicians) and FLJ (internal medicine physician) developed a checklist to describe adherence to the application of the *Guidelines for the care of patients in the region of the Americas - Second edition of the Pan American Health Organization* (4) and the Technical Health Standard for the Comprehensive Care of Patients with Dengue in Peru, 2024 (6). With both, the relevance of the initial diagnosis, severity classification, hydration (bolus and infusion) and monitoring from the health centers/posts until passing through the hospital were evaluated. Dengue (DENV) was defined as WHO: “*Dengue (break-bone fever) is a viral infection that spreads from mosquitoes to people. It is more common in tropical and subtropical climates. Most people who get dengue will not have symptoms. But for those who do, the most common symptoms are high fever, headache, body aches, nausea, and rash. Most will get better in 1–2 weeks. Some people develop severe dengue and need care in a hospital “*(7).

The algorithms followed were:

#### *A.* Dengue case algorithm with warning signs

1. Start hydration with crystalloid solution (Hartman or 0.9% saline solution): 10cc/kg to pass in one hour, up to 3 loads; If there is no clinical improvement change to the severe dengue algorithm.
2. Gradual reduction of hydration to 5-7 ml/kg/h for 2-4 hours and hourly monitoring.
3. Reevaluation of the patient. If clinical improvement is evident and urine output is ≥ 1 ml/kg/h, reduction of hydration to 3-5 ml/kg/h for 2-4 hours and hourly monitoring.
4. Reevaluation. If clinical improvement is evident and urine output is ≥ 1 ml/kg/h, reduce hydration to 2-4 ml/kg/h and continue for 24-48 hours and hourly monitoring.

#### *B.* Severe Dengue Case Algorithm

1. Immediate bolus administration of crystalloid solution (Hartman or 0.9% normal saline) 20 ml/kg in 15 to 30 minutes.
2. Reevaluation. If signs of shock disappear, decrease hydration to 10 ml/kg/h for 1-2 hours and hourly monitoring.
3. Reevaluation. If the evolution is satisfactory, decrease hydration to 5-7 ml/kg/h for 4-6 hours and hourly monitoring.
4. Reevaluation. If the evolution is satisfactory, reduction of hydration to 3-5 ml/kg/h for 2-4 hours and hourly monitoring.
5. If evolution is satisfactory, reduce fluids to 2-4 ml/kg/h for 24-48 hours.

In addition, the following were measured: Presence of the referral form from the healthcare center (presence and adequate completion), time elapsed from the reference from the health center to arrival at the hospital, and times spent in each service.

Two aspects were determined: 1. Appropriate initial diagnosis and 2. Appropriate management. Each researcher was assigned a defined number of records. They were previously trained by SCR and AMR.

*Adequate initial diagnosis* was defined if the diagnosis that appeared on the reference, triage and observation/Uviclin sheet coincided with the investigator’s opinion: DWWS, DWS, SD.

*Adequate management* was defined if the indications in the evolution or monitoring sheets coincided with the researcher’s evaluation in relation to parenteral fluid therapy, the frequency of monitoring vital functions (mean arterial pressure, diuresis) and the indication for blood products (all should be met).

For the evaluation of diagnosis and management of Dengue, confirmed and probable cases of dengue were included, as well as those ruled out since the outbreak scenario and the concept of case management were considered. If the written information was not found in the clinical history, it was considered indeterminate.

### Statistical Analysis

Descriptive analysis with absolute and relative frequencies for categorical variables and measures of central tendency and dispersion for numerical variables were estimated. The association between severity and mortality and adequate initial diagnosis and appropriate management was explored. Chi ^2^ and Fisher’s test were used for categorical tests and Wilcoxon/Student’s t test for numerical tests according to normality. A multivariate analysis was performed between mortality and diagnostic classification, sociodemographic and appropriate management classification with p <0.1 in the bivariate. Logistic regression with backward approximation was used.

### Ethical aspects

The Ethics Committee of the Teaching and Research Unit at the Hospital de la Amistad, Peru-Korea Santa Rosa, approved the study protocol. We delivered a final report to the hospital’s general management and shared the results with the authorities and the hospital’s dengue team.

## Results

The Minister of Health’s Epidemiology Surveillance System (ESS) reported 208 cases of DENV in 2022; between weeks 18 and 25, in 2023, they reported 731 cases, for a total of 939. We evaluated 141 medical records.

### General characteristics of these cases

Sixty-six percent of the medical records were from women, almost 67/141 (47.51%) were previously in a healthcare center or hospital other than Hospital Peru-Corea Santa Rosa, 31.20% had between 1-4 hours of delay in their referral, 56.73% were between 1-4 hours in triage, 76.60% had dengue with alarm signs, and 32% were in UVICLIN and deceases 18.44% (see table 1).

**Table 1:**
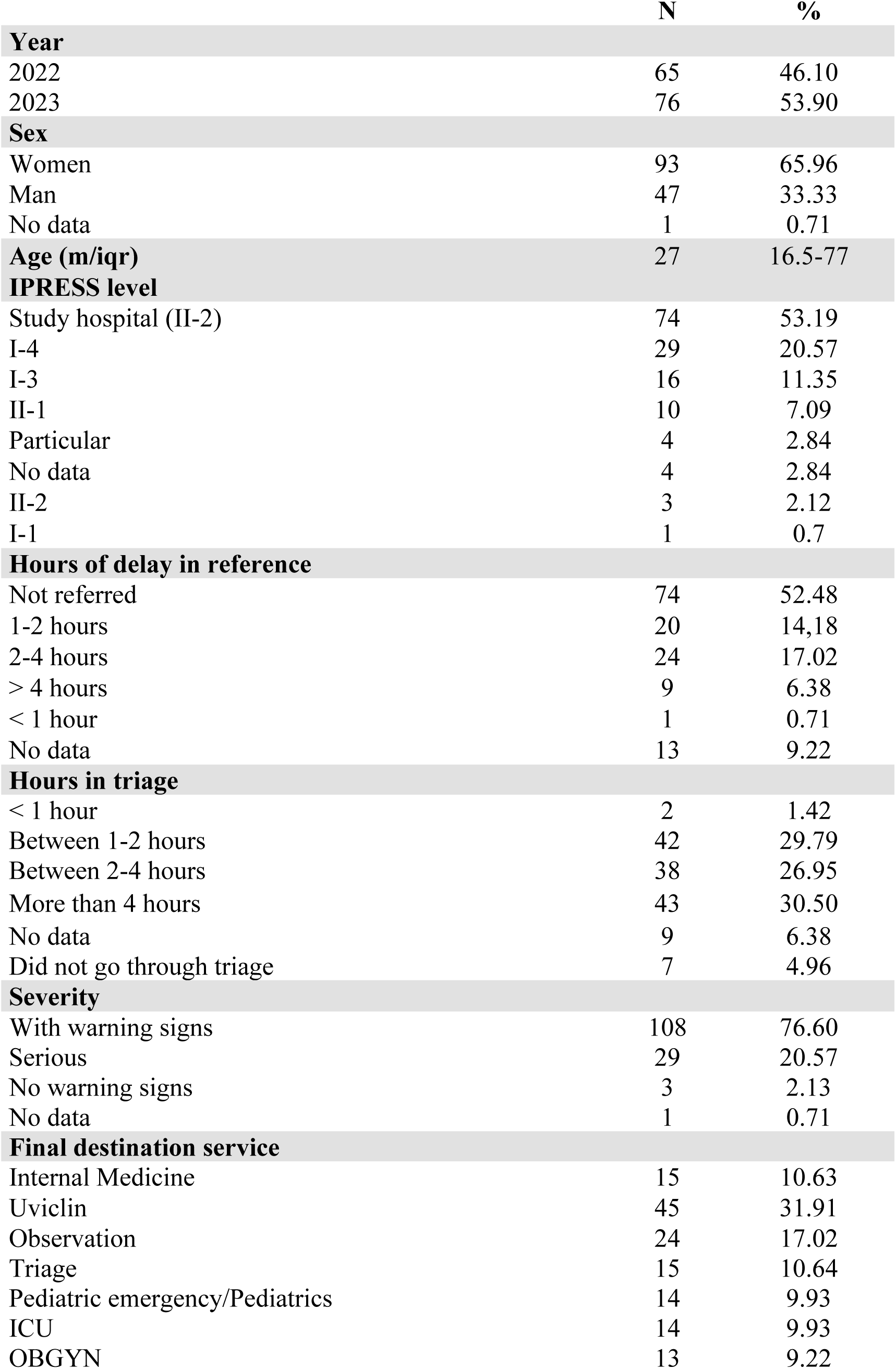
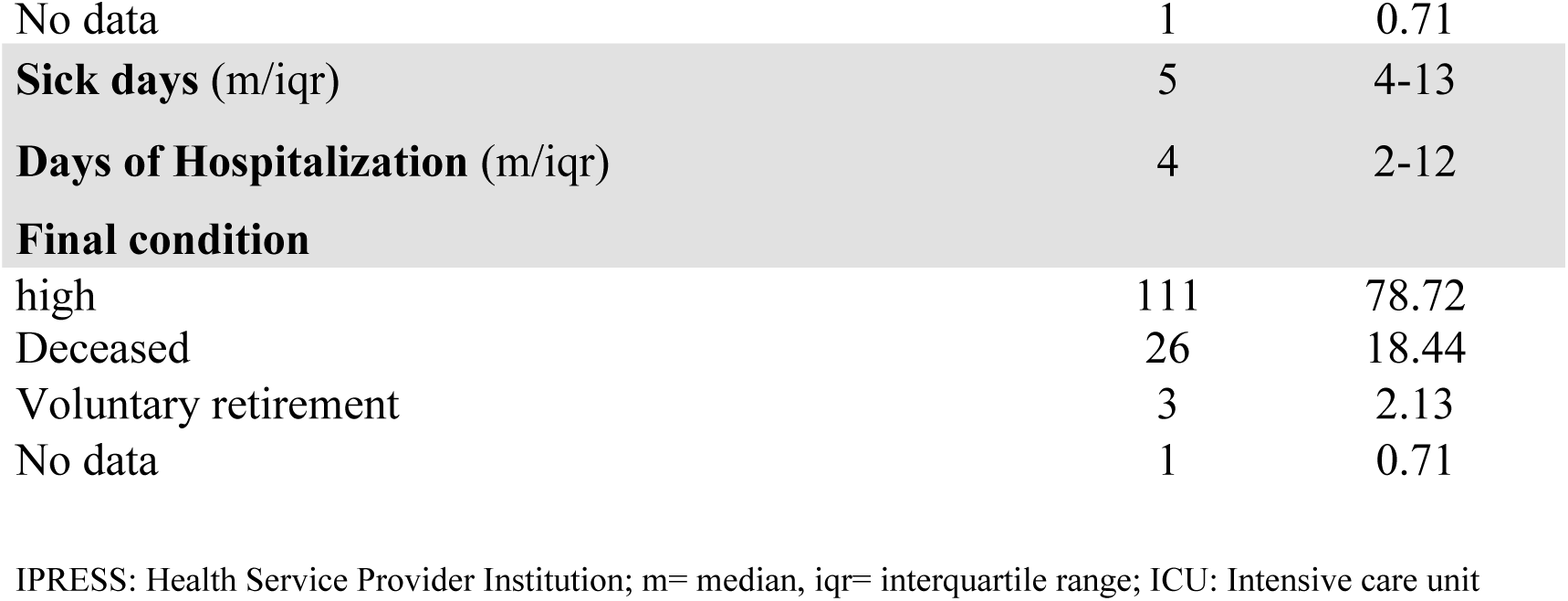
General characteristics of the total number of services (n=141)

When considering the classification of severity or treatment, at healthcare facilities outside of the study site, triage and Uviclin/observation, there were a frequency of at least one error in 91.5%, 88.5% and 52.6%, respectively (see table 2).

**Table 2:** Frequency of errors in diagnosis and treatment of Dengue by service (n=141), 2022-2023 period.

| Healthcare center/Hospital<br>(n=67) |  |  | Triage*<br>(n=134) |  |  | Observation/Uviclin//Hospitalization**<br>(n=114) |  |  |
| --- | --- | --- | --- | --- | --- | --- | --- | --- |
|  | N | % |  | N | % |  | N | % |
| **Diagnostic error** |  |  | **Diagnostic error** |  |  | **Diagnostic error** |  |  |
| Correct diagnosis | 35 | 52.23 | Correct diagnosis | 92 | 68.65 | Correct diagnosis | 95 | 69.6 |
| Incorrect diagnosis | 24 | 35.82 | Incorrect diagnosis | 41 | 30.59 | Incorrect diagnosis | 17 | 10.1 |
| Indeterminate | 8 | 11.94 | Indeterminate | 1 | 0.74 | Indeterminate | 2 | 1.2 |
| **Hydration** |  |  | **Hydration** |  |  | **Hydration** |  |  |
| Adequate | 24 | 35.82 | Adequate | 31 | 23.14 | Adequate | 62 | 54.3 |
| Not suitable | 36 | 53.73 | Not suitable | 103 | 76.86 | Not suitable | 50 | 45.6 |
| Indeterminate | 7 | 10.44 | Indeterminate | 0 | 0 | Indeterminate | 2 | 1.7 |
| **Urinary flow measurement** |  |  | **Urinary flow measurement** |  |  | **Urinary flow measurement** |  |  |
| Adequate | 11 | 16.41 | Adequate | 61 | 45.5 | Adequate | 95 | 83.3 |
| Not suitable | 49 | 73.13 | Not suitable | 72 | 53.7 | Not suitable | 17 | 14.9 |
| Indeterminate | 5 | 7.6 | Indeterminate | 1 | 0.7 | Indeterminate | 2 | 1.7 |
| **MAP control** |  |  | **MAP control** |  |  | **MAP control** |  |  |
| It was recorded | 41 | 61.19 | It was recorded | 89 | 66.41 | It was recorded | 99 | 86.8 |
| It was not recorded | 19 | 28.35 | It was not recorded | 45 | 33.58 | It was not recorded | 13 | 11.4 |
| Indeterminate | 7 | 10.44 | Indeterminate | 0 | 0 | Indeterminate | 2 | 1.7 |
| **platelet transfusion** |  |  | **platelet transfusion** |  |  | **platelet transfusion** |  |  |
| No | 67 | 100.0 | Yes | 2 | 1.4 | Yes | 9 | 7.8 |
| Yes | 0 | 0 | No | 132 | 98.5 | No/no data | 105 | 92.1 |
| **at least one mistake** |  |  | **at least one mistake** |  |  | **at least one mistake** |  |  |
| Yes | 56 | 83.58 | Yes | 119 | 88.80 | Yes | 63 | 55.2 |
| No | 3 | 4.47 | No | 15 | 11.19 | No | 48 | 42.1 |
| No data | 7 | 10.44 | No data | 0 | 0 | No data | 3 | 2.6 |

MAP: mean arterial pressure; \*: Triage: medicine, pediatrics and gynecology; \*\*: medicine, pediatrics and gynecology; only Uviclin has patients exclusively from this area.

In order to explore the association between sociodemographic features, hospital care variables and severity/mortality, voluntary withdrawals were considered alive until discharge and the observation of “missing data” was eliminated for the final outcome (n =140). Likewise, only those admitted with DWS and SD were considered (n=137). When comparing the presence of at least one error in the healthcare center, triage, observation/Uviclin, in all the three areas simultaneously and in the entire process, no differences were found between patients with DWS and SD: p=0.991, p= 0.136, p= 0.263, p=0.906 and p=0.249, respectively (see Table 3). In the exploratory multivariate analysis, only the severity of the disease was associated with mortality (OR 318.4, 95% IC [33.8-2996], p<0.001), other variable like age, year [2022-2023], not be referred, at least one error in the process were not significantly associated with mortality (see table 4).

**Table 3:** Bivariate analysis between care characteristics and Mortality (n=137)

| Time and attention management | Alive |  | Deceased |  |  |
| --- | --- | --- | --- | --- | --- |
|  | M | IQR | M | IQR | p value* |
| Days of illness | 4 | 4-13 | 3 | 4-8 | 0.842 |
| Hospitalization days | 3 | 4-12 | 3 | 0.29-9 | 0.359 |
| Age | 26 | 14.5-71 | 51 | 22-67 | 0.0031 |
|  | N | % | N | % | p** value |
| Sex |  |  |  |  |  |
| Man | 36 | 76.60 | 11 | 23.40 | 0.232 |
| Women | 75 | 83.33 | 15 | 16.67 |  |
| IPRESS Type |  |  |  |  |  |
| Study site | 55 | 75.34 | 18 | 24.66 | 0.083 |
| Others | 56 | 87.50 | 8 | 12.5 |  |
| Year of illness |  |  |  |  |  |
| 2022 | 58 | 90.63 | 6 | 9.38 | 0.0073 |
| 2023 | 53 | 72.60 | 20 | 27.40 |  |
| At least 01 error in HC |  |  |  |  |  |
| at least one mistake | 47 | 87.04 | 7 | 12.96 | 0.660 |
| No mistake | 4 | 80.0 | 1 | 20.0 |  |
| At least 01 error in Triage |  |  |  |  |  |
| at least one mistake | 91 | 79.13 | 24 | 20.86 | 0.189 |
| No mistake | 14 | 93.33 | 1 | 6.60 |  |
| At least 1 error in Observation /Uviclin |  |  |  |  |  |
| at least one mistake | 40 | 78.43 | 11 | 21.57 | 0.604 |
| No mistake | 38 | 82.61 | 8 | 17.39 |  |
| At least 1 error in the study site |  |  |  |  |  |
| Yes | 96 | 80.67 | 23 | 19.33 | 0.788 |
| No | 15 | 83.33 | 3 | 16.67 |  |
| At least 1 error in the entire process |  |  |  |  |  |
| Yes | 105 | 82.03 | 23 | 17.97 | 0.173 |
| No | 5 | 62.50 | 3 | 37.50 |  |
| Dengue severity |  |  |  |  |  |
| Dengue with warning signs | 105 | 97.22 | 3 | 2.78 | <0.001 |
| Severe Dengue | 6 | 20.69 | 23 | 79.31 |  |

M= median; IQR: Interquartile range; DWS: Dengue with warning signs; SD: Severe dengue; HC: healthcare center; \* Kruskal wallis test; \*\* chi 2

**Table 4:** Multivariate Analysis of the final model.

| Variables included | adjusted |  |  |
| --- | --- | --- | --- |
|  | OR | 95% CI | p value |
| Age | 0.95 | 0.91-0.99 | 0.094 |
| Year 2023 vs 2022 | 2.15 | 0.37-12.32 | 0.383 |
| Not be referred | 6.15 | 0.58-64.76 | 0.130 |
| At least 01 error in the process | 0.44 | 0.01-11.90 | 0.631 |
| Dengue severity | 318.4 | 33.8-2996 | <0.001 |

R<sup>2</sup>= 0.663

## Discussion

The study’s results indicate a high frequency of diagnostic and initial management errors in the pre-hospital and hospital phases. In both triage and the healthcare center, 8/10 treated patients experienced at least one error or omission during the process; in observation/Uviclin, this error occurred in 5/10 patients. Similarly, the healthcare center and triage incorrectly classified 1/3 of the patients based on their severity, which improved under observation. Only 3/10 patients in triage and 2/10 patients in the healthcare center, respectively, received adequate hydration.

The results may show bias in healthcare staff documentation because they may not have enough time to record every detail. Another possibility is a lack of standardized formats set up by the new National Technical Standard (5). This reality is often evident in scenarios of patient overload, epidemics, and care provided by front-line personnel with limited experience. A recent example was the COVID-19 pandemic, which made this type of inconvenience clear (15). On the other hand, a lack of adherence to the CPG recommendations is common not only in our country but in other realities and infectious pathologies such as COVID-19 (16).

Evidence suggests that the region has conducted training. Experts conducted four training sessions between 2023 and 2024: the first for 150 primary care health professionals (17), and the second for personnel with similar characteristics in the Morropon-Huancabamba Health Sub-Region (18). The Regional Directorate of Health Piura trained more than 1,200 nurses from the Ministry of Health (MINSA), Social Security (Essalud), Police Forces Health Facilities (Sanidad), and other healthcare establishments in a 12-hour workshop course in October 2023 (19, 20). However, it is crucial for any strategy to not only provide training for the personnel in charge but also to measure the baseline (knowledge, attitudes, and practices) (8) (10) and determine the impact afterwards. In this scenario, quasi-experimental studies are an attractive model and an especially useful tool (21). In an 18-month study in India by Balkrishnan et al., in patients with DWS, pre- and post-intervention measurements were carried out, determining that the educational program had an impact on improving the admission and classification criteria, monitoring of vital signs, correct use of bolus fluids, fluid therapy in titration, hematocrit monitoring, platelet transfusion when indicated, and discharge criteria. Also, mortality decreased from 7.1% to zero. The median hospital stay decreased by one day (22). An attractive possibility is to describe the before and after evaluations conducted on doctors in Piura in 2023 (20). Qualitative research is another unexplored scenario that is potentially helpful in exploring the perceptions, attitudes, and limitations of human resources. Quantitative measurement of adherence to a guideline is insufficient to explore these scenarios (23).

The Peruvian Health Ministry Directorate for the Prevention and Control of Metaxenic and Zoonotic Diseases developed a strategy to measure adherence to the National Technical Standard’s recommendations (5). According to their findings, in Peru, adherence was 70.5% (249 evaluations), with 51.27% in treatment and 73.76% in diagnosis. After reviewing 33 records, global adherence was 68.22% in Piura, 73.74% in diagnosis, and 49.19% in treatment, ranking ninth out of 13 regions (24). These figures are more encouraging; however, the evaluation of compliance should focus on the correct completion of forms and sociodemographic data rather than clinical aspects. We believe that our results can provide input for metaxenic strategies and other managers looking to improve training, infrastructure, and political commitment to addressing dengue. In the case of the Peruvian Social Security Health System (ESSALUD), we audited 42 clinical records as of March 2024 from cities Piura, Lambayeque, La Libertad, Ancash, and Ica. We found that the frequencies of notification to epidemiology surveillance, recording of the date of onset of symptoms, observation/hospitalization, parenteral hydration, recording of vital functions, and request for at least one blood count were 62%, 83%, 83%, 76%, 71%, and 91%, respectively (25). The National Health Ministry Health System made similar observations.

Even in patients with DWS, the high mortality rate of 18% is striking. Other probable causes include related comorbidities (which were not measured in this study) and limitations in diagnosis and case management. In a recent systematic review of our country, out of 17 publications between 1993 and 2023, including 10 regions of Peru and 2310 patients, mortality was 1.73% (26). The significant variation in mortality could potentially be attributed to our study’s bias, as we exclusively included patients from Piura, our non-random sampling method, and the severity of our patients (20.7% of whom had severe dengue). However, in the exploratory multivariate analysis, the presence of at least one error in the hospital, in the entire process, and in each of the areas, as well as other management problems, was not ultimately associated with mortality. This could be attributed to biases in the sample size (141/158) and sample design (consecutive), as well as the challenges of applying a retrospective checklist, which, despite training, may have measurement biases. Only dengue severity was associated with higher mortality.

There has been serious criticism of the efficiency of the reference and counter-reference systems in Peru. The referral process took more than 2 hours for more than 60% of patients (27). According to the regional government, the hospital where the study took place has approved the formation of a counter-referral team, but its implementation is still under development (28). Meanwhile, over 30% stayed in triage for over four hours. Understaffing and insufficient areas of attention are possible explanations. The current National Technical Standard mandates that in DWS, medical care should begin in less than 30 minutes, with immediate intravenous hydration; in SD, immediate attention is needed (5). This study has found many challenges in implementing these measures.

Regarding the observation/Uviclin area’s findings (55% of “at least one error”), we could attribute these findings to time constraints, patient overload, burnout, unnecessary form filling (National Insurance/discharge procedures), and possibly a lack of organization. This area requires monitoring with practical protocols. It is also imperative to verify the proper operation of equipment and supplies.

Theoretically, hospitalized first-level health care facilities (I-3 and I-4) can handle dengue diagnosis and alarm signs by providing an immediate response based on hydration and vital sign monitoring (blood pressure, heart rate, respiratory rate, temperature, oxygen saturation, water and electrolyte balance, etc.), which forms the foundation of effective dengue management. These facilities typically have a basic laboratory that works between 12 and 24 hours for monitoring with a complete blood count, although dengue management is still clinical in the Americas, according to the WHO (29). Again, human resource training (3, 4), as well as a lack of clear and socialized processes, could explain this reality.

One of the study’s limitations was that it did not meet the required sample size (n = 159 medical records). Some of the reasons for this were the inability to find the clinical history, incomplete data in the follow-up, and a lack of digitization of data and processes. However, the limitations prevented a random sampling process. Likewise, the 2023 cases were limited to weeks 18–25 (the peak of the outbreak). However, we believe that these results can serve to support any effort in the implementation of the electronic health record to improve information management (30). The Ministry of Health and the regional governments still have unresolved issues.

In conclusion, we found a high frequency of errors in the diagnosis of severity and in the treatment of patients with DWS and SD in the pre-hospital and hospital phases, but the sample size was insufficient to determine whether this finding had an impact on mortality.

## Data Availability

The database has been included in the supplementary information.

## Acknowledgments

to the Hospital authorities for the permissions provided for data access.

## REFERENCES

1. Pan American Health Organization. PLISA Health Information Platform for the Americas. Available at: https://www3.paho.org/data/index.php/en/mnu-topics/indicadores-dengue-en/dengue-nacional-en/252-dengue-pais-ano-en.html. Access date: February 12, 2024.

2. Ministry of Health. National Center for Epidemiology, disease control and prevention. Dengue Situation Room. Available at: https://www.dge.gob.pe/sala-situacional-dengue/diaria/. Access date: February 12, 2024.

3. World health organization: Dengue and severe dengue. Available at: https://www.who.int/es/news-room/fact-sheets/detail/dengue-and-severe-dengue. Access date: August 17, 2022.

4. Pan American Health Organization. Dengue: Guidelines for the care of patients in the Region of the Americas (2nd Edition). Available at: https://www.paho.org/es/documentos/dengue-guias-para-atencion-enfermos-region-americas-2a-edicion. Access date: May 22, 2023.

5. Ministry of Health. Ministerial Resolution. 082-2024-MINSA. Dengue Prevention and Control Plan 2024. Available at: https://cdn.www.gob.pe/uploads/document/file/5813240/5156456-resolucion-ministerial-n-082-2024.pdf. Access date: 3/26/2024.

6. Ministry of Health. Ministerial Resolution 175-2024/MINSA. Technical Health Standard for the comprehensive care of patients with Dengue in Peru. Available at: https://cdn.www.gob.pe/uploads/document/file/6007546/5323501-rm-175-2024-minsa-y-nts-211-dgiesp.pdf?v=1709834791. Access date: 03/10/24.

7. Organización Mundial de la Salud. Dengue y Dengue Grave. Disponible en: https://www.who.int/es/news-room/fact-sheets/detail/dengue-and-severe-dengue. Access date: 18/06/2024.

8. Koonisetty KS, Aghamohammadi N, Urmi T, Yavaşoglu Sİ, Rahman MS, Nandy R, et.al. Assessment of Knowledge, Attitudes, and Practices Regarding Dengue among Physicians: A Web-Based Cross-Sectional Survey. Behav Sci (Basel). 2021 Jul 21;11(8):105. doi:10.3390/bs11080105.

9. Laoprasopwattana K, Khantee P, Saelim K, Geater A. Mortality Rates of Severe Dengue Viral Infection Before and After Implementation of a Revised Guideline for Severe Dengue. Pediatr Infect Dis J. 2022 Mar 1;41(3):211–216. doi: 10.1097/INF.0000000000003411.

10. León F, Inga-Chero J, Mendoza-Farro N, Montoya A, Dioses K, Cavalcanti S, Moyano L. Characteristics and most frequent errors in the diagnosis and treatment of dengue in a hospital in northern Peru, 2022. An Fac med. 2023; 84(2):210–212. DOI: 10.15381/anales.v84i2.25494.

11. Rodríguez J, Lastre G, Camargo J, Fuentes G, Bermejo J, Nieto V. Patient adherence to the Dengue Clinical Practice Guideline (GPC_Dengue) in a Clinic in Barranquilla (Atl, Col). Cienc. Innov. Salud. 2015; 3 (2):23 – 30.

12. Valderrama Ardila M, Velasco M, Molina Lopez A, Perea Ayala M, Noguera Quiñones D, Rodriguez Arenas D, Velásquez PA. Evaluation of the management of pediatric patients diagnosed with dengue in a medium-complexity children’s clinic, according to the new WHO guideline. Rev. Col. Salud libre. 2011; 9:33–44.

13. Ministry of Health. Population statistics. Available at: https://www.minsa.gob.pe/reunis/data/poblacion_estimada.asp. Access date: 04/18/24.

14. Peru-Korea Santa Rosa Friendship Hospital II-2. Available at: https://www.gob.pe/hsantarosa. Access date: September 23, 2022.

15. Lake EA, Demissie BW, Gebeyehu NA, Wassie AY, Gelaw KA, Azeze GA. Knowledge, attitude and practice towards COVID-19 among health professionals in Ethiopia: A systematic review and meta-analysis. PLoS One. 2021 Feb 19;16(2): e0247204. doi: 10.1371/journal.pone.0247204.

16. Carrera-Acosta Lourdes, Salvador-Salvador Stefany, Torre-Maraví Gloria Edith. Evaluation of adherence to Clinical Practice Guides in the Social Security of Peru. Rev. Cuerpo Med. HNAAA 2021. 14(4): 430–431. 10.35434/rcmhnaaa.2021.144.1355.

17. Piura Regional Health Directorate. Health personnel training to strengthen health interventions against dengue. Press release. Available at: https://www.gob.pe/institucion/regionpiura-diresa/noticias/752982-capacitan-al-personal-de-salud-para-fortalecer-las-intervenciones-sanitarias-contra-el-dengue. Access date: May 30, 2023.

18. Piura Regional Government. Strengthening the capacities of health personnel for better care of patients with dengue in Alto Piura. Press release. https://www.gob.pe/institucion/regionpiura-dsrsmh/noticias/768037-fortalecen-capacidades-de-personal-de-salud-para-una-mejor-atencion-de-pacientes-con-dengue-en-el-alto-piura.

19. Ministry of Health. Piura: doctors are trained to care for patients affected by dengue. Available at: https://www.gob.pe/institucion/minsa/noticias/843478-piura-medicos-son-capacitados-para-atender-a-pacientes-afectados-por-dengue. Access date: 03/26/24.

20. Ministry of Health. Piura Regional Health Directorate. More than 1,200 nurses are trained to deal with dengue in the Piura region. Available at: https://www.gob.pe/institucion/regionpiura-diresa/noticias/900660-mil-200-enfermeras-son-capacitadas-para-enfrentar-al-dengue-en-la-region-piura. Access date: 3/26/24.

21. UNICEF. Methodological synthesis. Synopsis of Impact Assessment N 8. Available at: https://www.unicef-irc.org/publications/pdf/MB8ES.pdf. Access date: June 6, 2023.

22. Balkrishnan P, Panda PK, Pandey RM, Biswas A, Aggarwal P, Vikram NK, et.al. Compliance of WHO Guideline on Dengue Management among Indian Patients: An Interventional Quality Improvement Study. J Assoc Physicians India. 2019 Apr;67(4):30–34. PMID: 31299835.

23. Wharton-Smith A, Green J, Loh EC, Gorrie A, Omar SFS, Bacchus L, Lum LCS. Using clinical practice guidelines to manage dengue: a qualitative study in a Malaysian hospital. BMC Infect Dis. 2019 Jan 11;19(1):45. doi:10.1186/s12879-019-3680-5.

24. Ministry of Health. Direction of prevention and control of metaxenic and zoonotic diseases. Evaluation of adherence to technical health standards. Available at: https://lookerstudio.google.com/u/0/reporting/695d47ab-cb34-48ee-b1dc-471925689515/page/xpKkD. Access date: 04/15/2024.

25. Evaluation of adherence to the “Clinical Practice Guide for the Care of Dengue Cases in Peru” in ESSALUD, as of 03/12/2024. Available at: https://ietsi.essalud.gob.pe/wp-content/uploads/2024/03/Reporte.pdf. Access date: 04/15/2024.

26. Ledesma Negreiros GC, Rodriguez Vásquez S, Valencia Hipólito JV. Clinical characteristics and epidemiological situation of dengue in Peru: A Systematic Review. Rev. Cuerpo Med. HNAAA. 2024; 17(1). Doi: 10.35434/rcmhnaaa.2024.171.2409

27. Llanos Zavalaga Luis Fernando, Orellana Vásquez Alberth Teófilo, Aguado Taquire Henry Francisco. Initial evaluation of the outpatient Reference and Counter-Reference System in the DIRIS Lima Norte, from the maternal and child centers. Rev Med Hered 2021. 32(2): 91-102. 10.20453/rmh.v32i2.3982

28. Piura Regional Government. Piura Regional Health Directorate. Peru-Korea Friendship Hospital Santa Rosa II-2. Formation of the References and Counter-References team. Directorial Resolution: N 602022/GOB.PIURA-DRSP-HAPCSR-II-2DIR-UDS. Available at: https://www.hsantarosa.gob.pe/PORTAL/RD_060.pdf. Access date: June 6, 2023.

29. Pan American Health Organization. Declaration of Alma-Ata. Washington DC: Pan American Health Organization; 2012. (Cited May 30, 2023). Available at: https://www.paho.org/hq/dmdocuments/2012/Alma-Ata-1978Declaracion.pdf.

30. Sampaio VS, Lopes R, Ozahata MC, Nakaya HI, Sousa E, Araújo JD, Bragatte MAS, Brito AF, Grespan RMZ, Capuani MLD, Domingues HH, Pellini ACG, Mateos SOG, Conde MTRP, Eudes Leal F, Sabino E, Simão M, Kalil J. Thinking outside the box: revisiting health surveillance based on medical records. Antimicrob Steward Health Epidemiol. 2023 Oct 24;3(1): e185. doi: 10.1017/ash.2023.451.

